# Differences in White Matter Structural Networks in Family Risk of Major Depressive Disorder and Suicidality: A Connectome Analysis

**DOI:** 10.1101/2023.09.07.23295211

**Authors:** Nora Clancy Kelsall, Yun Wang, Marc J Gameroff, Jiook Cha, Jonathan Posner, Ardesheer Talati, Myrna M. Weissman, Milenna Tamara van Dijk

## Abstract

**Background:** Depression and suicide are leading global causes of disability and death and are highly familial. Family and individual history of depression are associated with neurobiological differences including decreased white matter connectivity; however, this has only been shown for individual regions. We use graph theory models to account for the network structure of the brain with high levels of specialization and integration and examine whether they differ by family history of depression or of suicidality within a three-generation longitudinal family study with well-characterized clinical histories.

**Methods:** Clinician interviews across three generations were used to classify family risk of depression and suicidality. Then, we created weighted network models using 108 cortical and subcortical regions of interest for 96 individuals using diffusion tensor imaging derived fiber tracts. Global and local summary measures (clustering coefficient, characteristic path length, and global and local efficiencies) and network-based statistics were utilized for group comparison of family history of depression and, separately, of suicidality, adjusted for personal psychopathology.

**Results:** Clustering coefficient (connectivity between neighboring regions) was lower in individuals at high family risk of depression and was associated with concurrent clinical symptoms. Network-based statistics showed hypoconnected subnetworks in individuals with high family risk of depression and of suicidality, after controlling for personal psychopathology. These subnetworks highlighted cortical-subcortical connections including between the superior frontal cortex, thalamus, precuneus, and putamen.

**Conclusions:** Family history of depression and of suicidality are associated with hypoconnectivity between subcortical and cortical regions, suggesting brain-wide impaired information processing, even in those personally unaffected.

## 1. Introduction

Mental illnesses including Major Depressive Disorder (MDD) are the single leading global cause of disability(1, 2), and over 700,000 people die by suicide each year (3). A consistently reported risk factor for development of depression is parental history of depression(4–7), which increases risk of depression two-to-five-fold, and is associated with abnormalities in social, cognitive and neurobiological structure and function, regardless of presence of personal psychopathology(5, 8–10). Depression in turn is a risk factor for suicidal thoughts, suicide attempts, and completed suicide(11). However, suicidality is an independent phenomenon and can occur in individuals without depression(12). Family history of suicide also increases an individual’s risk of suicide, beyond the effects of family history of depression(13, 14). However, little is known about the neurobiological differences associated with family risk of depression and suicidality. Elucidating the effects of family risk of depression and suicidality on brain structure and function could ultimately allow for treatment and prevention strategies.

Neuroimaging studies commonly investigate neurobiological differences related to depression or suicidality using isolated brain regions and individual connections in a mass univariate approach. They have associated MDD with prefrontal and subcortical volume loss(15, 16) including differential hippocampal subfield volume loss(17–19) and decreased limbic-prefrontal and thalamic-prefrontal white matter connectivity(20–22). Similarly, (unaffected) individuals with family history of MDD showed cortical thinning, decreased putamen and hippocampal volumes(23–26), and decreased integrity in cortical-subcortical white matter tracts such as the uncinate fasciculus and cingulum(27). Furthermore, gray matter microstructure in the dentate gyrus, a subfield of the hippocampus, was decreased in individuals at high family risk of depression, regardless of personal depression status. Lower microstructure predicted future but not past or current symptoms(28). Individuals with a personal history of suicidality have decreased volume in regions such as the hippocampus, amygdala, putamen, and thalamus (29–31).

The brain is a network that is both highly integrated and highly specialized and segregated. In the last decade network science and graph theory, including Network Based Statistics (NBS), has become increasingly popular to study how the brain performs complex cognitive processes and balances integration and segregation(32, 33). Graph theory represents brain regions as nodes in a graph to create a single model representing the entire brain. This approach allows for calculation of summary measures that characterize how well the brain is integrated as a whole, so-called global measures, and how well each region is connected to its neighbors, so-called local measures. NBS analysis uses the graph model to identify subnetworks that differ in connectivity between populations of interest, rather than investigating one connection at a time. The use of bootstrapping to evaluate subnetwork significance circumvents the common issue of multiple comparisons when evaluating individual connections. Thus, graph theory allows us to investigate brain differences between populations of interest as an integrated and specialized network, instead of evaluating specific pathways or connections “out-of-context”.

Studies using NBS and graph theory identified subnetworks that are associated with MDD (34–36) as well as with suicidality(37–39). Nonetheless, despite growing research in this area, to our knowledge, graph theory and NBS have not been used to investigate whether family risk of depression or suicidality is associated with brain-wide and subnetwork hypoconnectivity.

We leverage a unique cohort of three generations at high and low risk of depression followed for up to 40 years which includes in depth clinician-based depression and suicidality diagnoses. We examine brain networks associated with family history of depression and of suicidality, using graph theory models to account for the high levels of neurological specialization and integration. Modeling white matter connectivity (measured by diffusion tensor imaging (DTI) tractography) as graphs, we calculated summary measures of the overall connectivity of the brain, with connectivity hypothesized to negatively correlate with family risk of MDD as well as family risk of suicidality. Using NBS methods, the models of brain connectivity were then analyzed to identify which specific subnetworks were hypoconnected.

## 2. Methods and Materials

### 2.1. Participants

Participants were drawn from a longitudinal high-risk study that began in the New Haven, CT metropolitan area in 1982. Probands (Generation 1:G1) and their offspring (Children: G2, and Grandchildren: G3) were followed up to 38 years across 7 waves of data collection. Detailed description of study procedures and recruitment can be found in(5). For this study, participants were included from the second and third generations of the study, who had clinical information on history of MDD and suicidality, and had MRI data collected. Spouses and children not biologically related to the original participants were excluded. Using these inclusion criteria, the analytic sample was n=97. All procedures were approved by the IRB of the New York State Psychiatric Institute and all participants provided written informed consent.

### 2.2 Exposure Classification

#### Risk of Depression

G1 consisted of two groups: 1) participants with a history of moderate to severe MDD seeking treatment at outpatient facilities, and 2) participants with no history of psychiatric illness, based on multiple Schedule for Affective Disorders and Schizophrenia (SADS) interviews (40). Subsequent generations of the families (G2, G3) were then categorized as high family risk of MDD or low family risk of MDD based on the MDD status of G1s.

For personal history of MDD, a Kiddie-SADS (K-SADS)(41) or SADS interview was completed at multiple waves including at the time of MRI by highly trained individuals who were blind to G1 depression history. Individuals were identified as having a personal history of MDD if they were diagnosed with MDD at or prior to any of these diagnostic interviews, prior to and including the time of the MRI.

#### Risk of Suicidality

Personal and family risk of suicidality was determined by the same (K)SADS interviews which included questions on presence and severity of suicidal ideation, as well as suicide attempts. Additionally, suicide completions were reported by family remembers. Suicidal ideation predicts future attempts, and thus was included to allow us to have a more complete understanding of individual and family suicidality(12). Suicidal ideation was considered moderate to severe by the clinician if the individual reported frequent thoughts of suicide including thoughts of a specific method of suicide, made a plan to attempt suicide, or made preparations for an attempt. An individual was considered as having a personal history of suicidality if they ever had reported moderate to severe suicidal ideation or at least one suicide attempt. A family was considered at high risk of suicidality if at least one individual in that family died by suicide, made a suicide attempt or had moderate to severe suicidal ideation. If a family was considered at high risk of suicidality, all family members were included in this category.

There were 96 participants included in this analysis. Based on the above classification, 46 were considered at low family risk and 50 were at high family risk of MDD. 59 individuals did not have a personal history of MDD and 37 individuals did have a history of MDD. Based on the family history of suicidality, 26 were considered at low family risk and 70 were at high family risk of suicidality. 80 individuals reported no suicidal ideation or mild ideation, while 16 reported a history of moderate to severe ideation or a past suicide attempt (Table 1).

**Table 1a.** Demographical information of participants on age, sex and exposure classification.

|  |  | Ages |  | Number of Subjects by Sex |  |  | Total<br>Number of<br>Subjects |
| --- | --- | --- | --- | --- | --- | --- | --- |
| | | Mean<br>(Standard Error) | t-value | Male | Female | $\chi^2$ | |
| Personal history<br>of MDD | Not present | 28.46 (1.723) | 3.99* | 32 | 27 | 0.00 | 59 |
|  | Present | 38.77 (2.074) |  | 20 | 17 |  | 37 |
| Family Risk of<br>MDD | Low | 29.93 (1.911) | 1.84 | 23 | 23 | 0.34 | 46 |
|  | High | 34.82 (2.032) |  | 29 | 21 |  | 50 |
| Personal history<br>of Suicidality | Not present | 32.13 (3.486) | 0.74 | 41 | 39 | 1.01 | 80 |
|  | Present | 35.03 (1.602) |  | 11 | 5 |  | 16 |
| Family risk of<br>Suicidality | Low | 35.76 (2.811) | -1.32 | 14 | 12 | 0.00 | 26 |
|  | High | 31.31 (1.627) |  | 38 | 32 |  | 70 |

\*p < 0.05

**Table 1b.** Participant overlap of family risk of suicidality and family risk of MDD.

|  | Low Family<br>Risk of<br>Suicidality | High Family<br>Risk of<br>Suicidality |
| --- | --- | --- |
| Low Family Risk<br>of MDD | n = 14 | n = 32 |
| High Family Risk<br>of MDD | n = 12 | n = 38 |

**Table 1c.** Participant overlap of personal history of suicidality and MDD.

|  | No personal history of suicidality | Personal history of suicidality |
| --- | --- | --- |
| No personal history of MDD | n = 57 | n = 2 |
| Personal history of MDD | n = 23 | n = 14 |

### 2.3 Assessments

In addition to (K)SADS to assess personal and family diagnoses for depression and suicidality, we selected symptom scales to assess associations between graph theory measures (at 50% consistency) and concurrent symptoms and 8-year follow-up symptoms. For symptoms measured at the time of scan Hamilton Anxiety (HAM-A) and Hamilton Depression (HAM-D)(42, 43) Rating Scales were available. For eight-year follow-up symptoms we selected the subscales for general depression and suicidality from the Inventory of Depression and Anxiety (IDAS)(44) since Hamilton scales were not used for assessments at that time.

#### 2.3. MRI Data Collection and Pre-Processing

Magnetic resonance imaging utilized a 3.0T General Electric (Milwaukee, Wisconsin) Signa HDx scanner with an 8-channel head coil as reported previously(26). DTI parameters included 70 axial contiguous 2.5mm slices; 1.72mm×1.72mm resolution; 128×128 matrix; repetition time=17,000msec; echo time=95msec; frequency direction = right/left; 42 diffusion orientations; and b-value=1250.

Using Freesurfer 6.0, T1-scans for each individual were separated into 108 different regions of interest (ROIs) based on the Desikan-Killany cortical parcellation atlas in combination with Freesurfer’s subcortical segmentation and the (para)hippocampal subfield segmentation “FS60” module (Supplemental Table 1 for all regions).

After visual inspection of images, DTI stream counts were calculated between each region by processing the images using MRtrix pipeline(45)(https://www.mrtrix.org/) as described previously(26). The pipeline includes denoising with random matrix theory(46), motion and eddy current correction, brain extraction, and bias field correction (N4 algorithm(47);N4ITK from Advanced Normalization Tools(48); ANTs). Probabilistic tractography based on second-order integration over fiber orientation distributions (iFOD2) with a target streamline count of 10 million(45) were used to construct the tractograms which were filtered using spherical-deconvolution informed filtering of tractograms (SIFT) with a streamline count target of 1 million. ROIs obtained from the T1-scan from the same individual were then used to create final connectivity matrices.

### 2.4. Network Construction

Using R4.2.1, stream counts between ROIs were normalized by individual total brain volume and stored in a 108×108 matrix for each subject (based on the 108 ROIs). Because we previously demonstrated that hippocampal subfield specific alterations predicted future depression(28), we included hippocampal subfields in our analyses.

R package Braingraph, Version 2.7.3(49) was used for graph construction with the 108 ROIs as network nodes and the normalized stream counts as the edge weights. The sample was divided into two groups by study objective (family risk of MDD, or family risk of suicidality), and connections were only retained if they were present in a higher percentage of the group than a given consistency threshold (e.g. 50%, 70% or 90%). For example, if the connection from the amygdala to the insula was present only in 35% of the subjects in a group, and the consistency threshold was 50%, the connection would not be included in the graph for any subject in that group(50) . This decreased the number of errant connections and allowed for maintenance of relatively consistent graph structure within each group.

### 2.5 Statistical Analysis

#### 2.5.1. Global and Local Graph Measure Analysis

Four global and local network measures were calculated for the graph of each person using Braingraph(49): clustering coefficient, weighted characteristic path length, weighted global efficiency, and weighted local efficiency, see Table 2 for definitions. These measures characterize brain integration as well as segregation and are commonly used in graph analysis including brain connectome analyses(51). Each of these measures were calculated for four different consistency thresholds, 0% (including all connections), 50%, 70%, 90%.

**Table 2.** List of definitions and equations for measures that were used for group comparison.

| Full Measure Name | Equation | Definition and interpretation |
| --- | --- | --- |
| Clustering coefficient | $C = \frac{1}{n} \sum_{i \in n} C_i = \frac{1}{n} \sum_{i \in N} \frac{2t_i}{k_i(k_i-1)}$<br>Where $C_i$ is the clustering coefficient of node $i$, $t_i$ is the number of triangles made (neighbors connected to each other), and $k_i$ is the number of neighbors | A measure of segregation, the fraction of a node's neighbors that are neighbors of each other, averaged across the network |
| Characteristic Path Length | $L = \frac{1}{n} \sum_{i \in n} L_i = \frac{1}{n} \sum_{i \in N} \frac{\sum_{j \in N, j \neq i} d_{ij}}{n-1}$<br>Where $L_i$ is the average distance between node $i$ and all other nodes, $d_{ij}$ is the number of steps between the given two nodes, and $n$ is the number of nodes total in the network | A measure of integration, the average shortest path in the network. More influenced by nodes that are relatively isolated from the network than global efficiency |
| Global Efficiency | $E = \frac{1}{n} \sum_{i \in n} E_i = \frac{1}{n} \sum_{i \in N} \frac{\sum_{j \in N, j \neq i} d_{ij}^{-1}}{n-1}$<br>Where $E_i$ is the efficiency of node $i$, $d_{ij}$ is the number of steps between the given two nodes, and $n$ is the number of nodes total in the network | The average inverse shortest path length in the network. Global efficiency is indicative of differing path lengths between regions and thus differences in efficiency, network integration, and information transmission (32, 53, 54). |
| Local Efficiency | $E_{loc} = \frac{1}{n} \sum_{i \in G} E_{glob}(G_i),$<br>Where $E_{glob}(G_i)$ is the global efficiency of the subgraph that contains only neighbors of node $i$. | Global efficiency computed only on node neighborhoods; thus, local efficiency of node $i$ characterizes how well information is exchanged by its neighbors when it is removed. It |
|  |  | is a measure of segregation, however unlike the clustering coefficient it does not directly evaluate number of connections between neighbors, and instead evaluates weighted path length within the subnetwork of a region's neighbors |

Generalized Estimating Equations (GEE) to account for family structure in the data were run to compare measures between groups based on family risk of MDD or family risk of suicidality. For analyses of family history of MDD and separately family history of suicidality, one primary model for each contrast and 4 additional sensitivity analyses were compared as detailed in Table 3 adjusting individually for personal history of MDD, family history of suicidality, personal history of suicidality, and all family and personal histories. All analyses included age and sex as covariates at each consistency threshold.

**Table 3.**
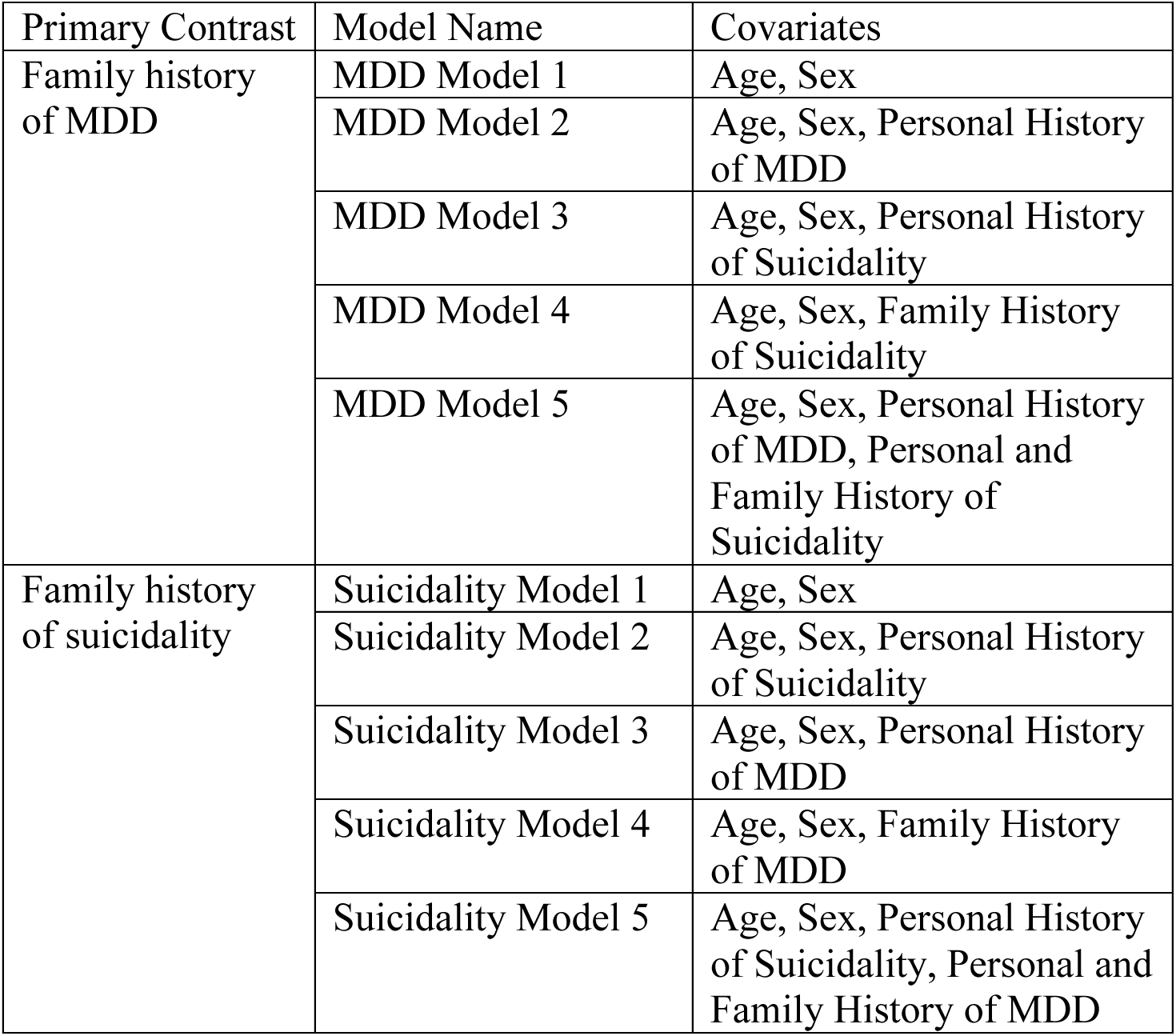
Included Covariates by Model.

| Primary Contrast | Model Name | Covariates |
| --- | --- | --- |
| Family history of MDD | MDD Model 1 | Age, Sex |
|  | MDD Model 2 | Age, Sex, Personal History of MDD |
|  | MDD Model 3 | Age, Sex, Personal History of Suicidality |
|  | MDD Model 4 | Age, Sex, Family History of Suicidality |
|  | MDD Model 5 | Age, Sex, Personal History of MDD, Personal and Family History of Suicidality |
| Family history of suicidality | Suicidality Model 1 | Age, Sex |
|  | Suicidality Model 2 | Age, Sex, Personal History of Suicidality |
|  | Suicidality Model 3 | Age, Sex, Personal History of MDD |
|  | Suicidality Model 4 | Age, Sex, Family History of MDD |
|  | Suicidality Model 5 | Age, Sex, Personal History of Suicidality, Personal and Family History of MDD |

Finally, we ran separate models between the graph measures that significantly differed between family risk groups at the 50% consistency threshold and concurrent symptoms (HAM-A and HAM-D) and follow-up symptoms (IDAS general depression and IDAS suicidality) adjusted for age and sex.

#### 2.5.2. Network Based Statistics Analysis

Braingraph was also used to run the Network Based Statistics analysis(52). For this analysis only the graphs maintaining connections consistent among 70% of the group were used as previously suggested(50) and connections were also removed if the mean stream count across all participants was less than 20 in order to maximize resolution, and minimize results based only on errant connections. The contrasts tested for hypoconnectivity as well as hyperconnectivity in groups of interest (i.e. High Family Risk > Low Family Risk, as well as Low Family Risk > High Family Risk). Covariates were adjusted in steps, as they were for the graph measure models, and described in Table 3.

NBS has four steps: 1) Test each connection between groups (e.g. High vs. Low family Risk of MDD). 2) Select an initial threshold to identify significant connections. For this study, 5 different initial thresholds were tested: 1×10^-6^, 1×10^-5^, 1×10^-4^, 5×10^-4^, 1×10^-3^. 3) Create subnetworks containing neighboring significant connections identified in step two. 4) Calculate a corrected p-value using permutation testing. We used n=5000 permutations to determine the corrected p-value for each subnetwork.

## 3. Results

### 3.1. Participants

Ninety-six participants from the cohort were included in the graph theory analysis (Table 1a for demographics). Thirty-eight participants were at high family risk of MDD and suicidality, 32 participants were at high family risk of suicidality, but low family risk of MDD, 12 were at high family risk of MDD, but low family risk of suicidality, and 14 were at low family risk of both suicidality and MDD (Table 1b). Correlation between high family risk of MDD and high family risk of suicidality was low (r(96) = 0.072, p = 0.484). On the other hand, only 2 out of 59 individuals without a personal history of MDD had a personal history of suicidality, whereas 14 out of 37 with a personal history of MDD also had a personal history of suicidality (Table 1c), and correlation between personal histories of MDD and suicidality was high (r(96)=0.450, p<.00001). Sex and head translation during DTI-scan were not significantly different by group for family risk of MDD, personal history of MDD, personal history of suicidality, or family risk of suicidality, and age was only significantly different by group for personal history of MDD. While depression and suicidality are related, the correlation is moderate, supporting the understanding of (family risk of) depression and (family risk of) suicidality as distinct psychological concepts to be studied here.

### 3.2. Global and Local Graph Measures

#### 3.2.1. Family risk of depression

The clustering coefficient was significantly lower in individuals at high compared to low family risk of depression across all consistency thresholds (p’s<0.005, Figure 1, Supplemental Table 2). A lower clustering coefficient indicates that regions are less likely to cluster together, and a lower proportion of a node’s (brain region’s) neighbors will be connected to each other on average. Clustering remained significantly decreased across the higher three consistency thresholds when adjusting for personal lifetime history of MDD, personal history of suicidality, family history of suicidality, and adjustment for all three (Supplemental Tables 3,4,5,6). At higher thresholds, because of the removal of more errant connections, groups have a more consistent structure, leading to more significant differences between groups.

**Figure 1.**
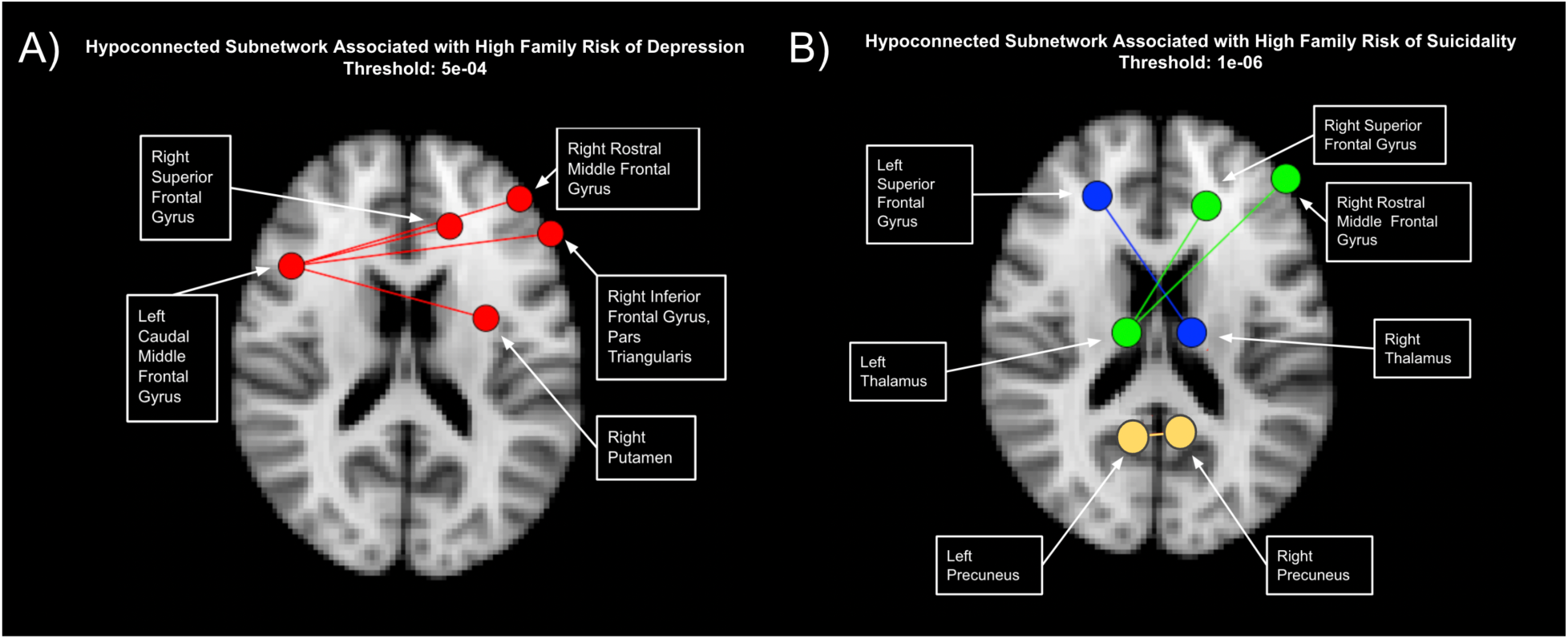
Association between family risk of MDD and brain summary graph measures at different thresholds. Connections were only retained in a graph if they were present in a higher percentage of the group than a given consistency threshold (e.g. 50%, 70% or 90%), leading to different estimations of summary measures. Clustering parameter was decreased in individuals with high family risk of MDD across all consistency thresholds. Local efficiency was only significantly decreased in individuals at high risk of MDD at the 50% threshold. (*** p-value < .005, ** p-value < .05)

Local efficiency was significantly lower in individuals at high family risk of depression, suggesting reduced robustness of connectivity of neighboring regions. However, this association was only significant at the 50%, and marginal at the 90% threshold suggesting reduced connectivity only for more consistent connections. This pattern remained the same after adjusting for all covariates (Supplemental Tables 3,4,5,6).

Global efficiency and characteristic path length were not associated with family risk of depression.

There was no significant interaction between family history of MDD and sex for characteristic path length, clustering coefficient, global efficiency, or local efficiency.

#### 3.2.2. Family risk of suicidality

High family risk of suicidality was associated with clustering coefficient but at the 90% threshold, and local efficiency was only significant at the 50% threshold, showing less consistent evidence for hypoconnectivity (Figure 2, Supplemental Table 7). The patterns remained significant across all covariate adjustments (Supplemental tables 8,9,10,11).

**Figure 2.**
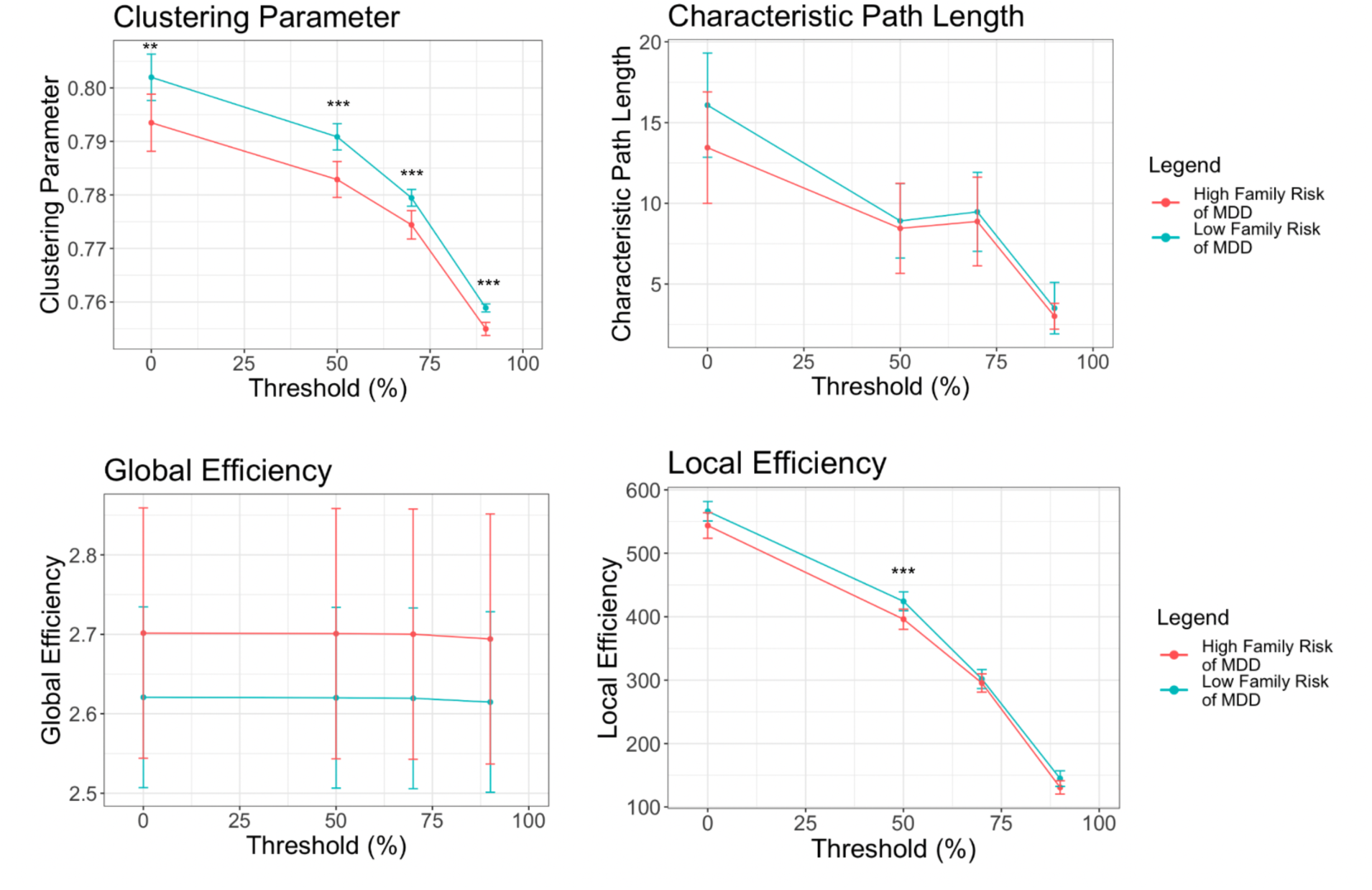
Association between family risk of suicidality and brain summary graph measures at different thresholds. Connections were only retained in a graph if they were present in a higher percentage of the group than a given consistency threshold (e.g. 50%, 70% or 90%), leading to different estimations of summary measures. Clustering parameter was decreased in individuals with high family risk of suicidality across at the 90% threshold. Local efficiency was significantly decreased in individuals at high risk of suicidality at the 50% threshold. (*** p-value < .005, ** p-value < .05)

Global efficiency and characteristic path length were not associated with family risk of suicidality. There was no significant interaction between family risk of suicidality and sex for characteristic path length, clustering coefficient, global efficiency, or local efficiency.

#### 3.2.3. Associations with Clinical Symptoms

We investigated whether the measures that significantly differed between familial risk groups (clustering coefficient and local efficiency) were associated with clinical symptoms at time of scan and 8-years post-scan.

Clustering coefficient (but not local efficiency) was associated with both depressive (st.beta=0.04, p=0.029) and anxiety (st.beta=-0.04, p=0.015) symptoms at the time of scan. Neither clustering coefficient nor local efficiency was associated with future symptoms measured by IDAS general depression and IDAS suicidality (ps>0.1).

### 3.3. NBS Analysis

For each NBS analysis, five different initial thresholds were tested, from 1×10^-6^ to 1×10^-3^. This initial threshold determines the significant threshold for connections to be included to identify subnetworks in the first step (see Methods). To illustrate, Supplemental Figure 1 compares two different initial thresholds: p = 1×10^-4^ and p = 5×10^-4^. The lower initial threshold is significantly more restrictive in which connections are included in subnetworks, and thus only identifies a four-region subnetwork, where-as the higher initial threshold identifies a subnetwork with 10 regions. For each comparison, the most restrictive threshold that contained a significantly hypoconnected subnetwork was used to present results. 5×10^-4^ is used to present results for family risk of depression, and 1×10^-6^ is used for family risk of suicidality, additional thresholds are shown in Supplemental Figures 2 and 3 for the reader’s interest. In the final step bootstrapping is used to establish the significance level of the entire subnetwork. In figures, subnetworks are displayed if the subnetwork has a final significance level below p = 0.05.

**Figure 3.**
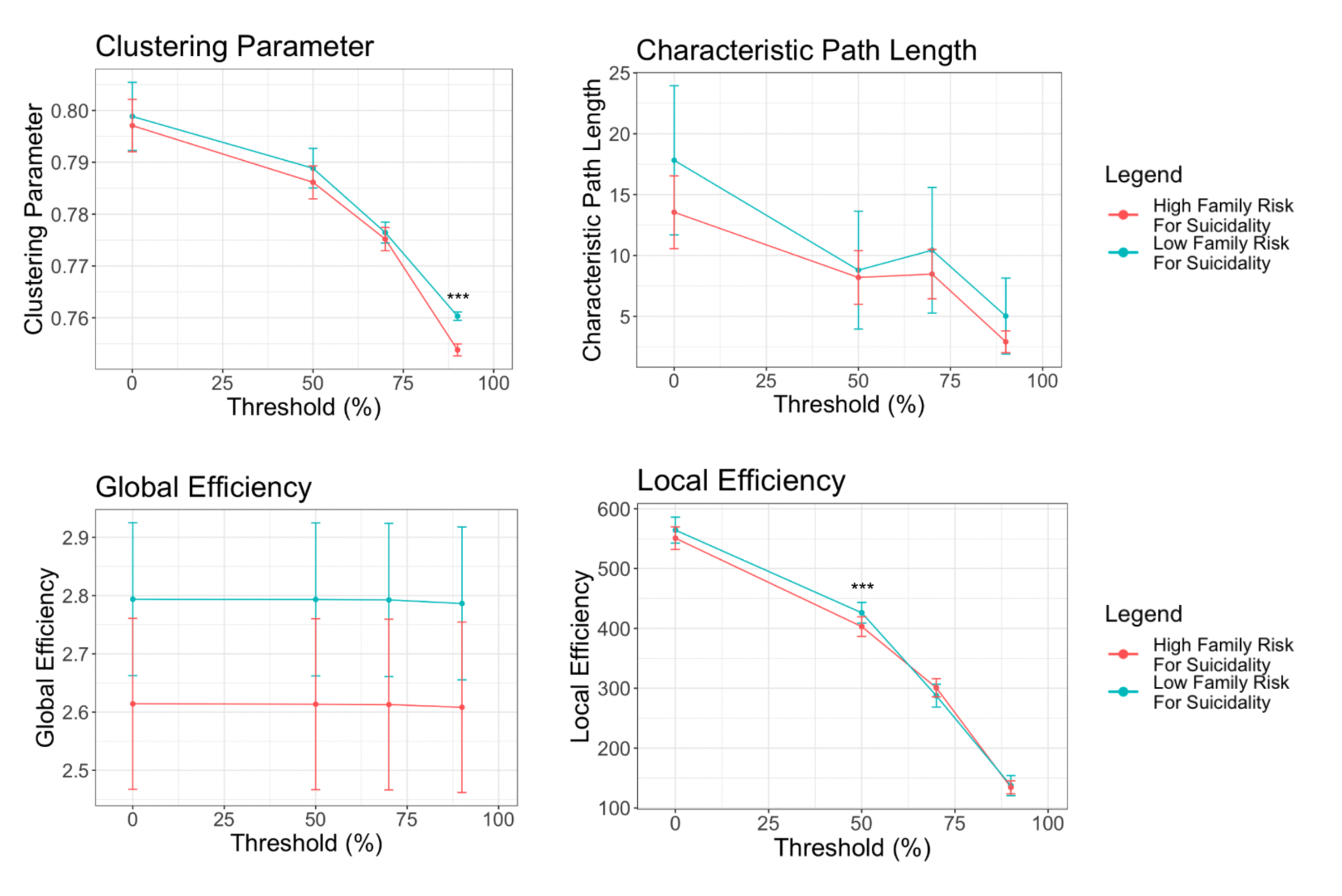
Comparison of networks associated with family risk of depression and suicidality. Subnetwork hypoconnected in individuals at A) high family risk of MDD includes the five regions: left caudal middle frontal cortex, right superior frontal cortex, right putamen, right rostral middle frontal cortex, and the right pars triangularis was significantly different (red network: p = 0.0108) after permutation testing between the two risk groups. and B) high family risk of suicidality. The 3 subnetworks that are significantly different (yellow network: p = 0.00040, blue network: p = 0.0046, and green network: p = 0.0046) contain 7 vertices and 4 edges. The yellow network includes right and left precuneus, the blue network includes the left superior frontal gyrus, and the green network includes the left thalamus, the right superior frontal gyrus, and the right rostral middle frontal gyrus.

#### 3.3.1. Family risk of MDD

A subnetwork including 5 regions was significantly hypoconnected in individuals with high family risk of depression (p=0.0108, Figure 1A) after adjusting for age and sex (Model 1). This subnetwork was consistent across higher initial thresholds. The subnetwork included the following regions: left caudal middle frontal gyrus, right superior frontal gyrus, right putamen, right rostral middle frontal gyrus, and the inferior frontal gyrus, pars triangularis. This association was robust to accounting for personal and family depression and suicidality covariates (for adjusted models 2-4 see Supplemental Results). For example, after adjusting for personal history of MDD, and personal and family history of suicidality (Model 5), the network was similar (p = 0.0212), however the putamen was not included until higher threshold subnetworks (Supplemental Figure 7). There were no subnetworks that were hyperconnected in individuals with high family risk of depression.

#### 3.3.2. Family Risk of Suicidality

Three subnetworks including 7 regions total were significantly hypoconnected in individuals with high family risk of suicidality (ps<0.01, Figure 1B) after adjusting for age and sex (Model 1). The first subnetwork (p=0.00040) contained three regions: the left thalamus, right rostral middle frontal cortex, and the right superior frontal gyrus. The second subnetwork contained two regions: right thalamus and the left superior frontal gyrus (p=0.0046), and the third subnetwork also contained two regions: the left precuneus and the right precuneus (p=0.0046). For further adjusted models 2-4 see Supplemental Results. After adjusting for personal history of suicidality, and personal and family history of MDD (Model 5), one subnetwork remained connecting the left thalamus and the right superior frontal gyrus. (p=0.0024; Supplemental Figure 11). Furthermore, after removing any individuals with personal history of suicidality, and adjusting for age and sex, the same subnetwork remained connecting the left thalamus and the right superior frontal gyrus in the remaining participants (n = 80, p=0.0014; Supplemental Figure 12). Thus, this connection was most robust to accounting for personal and family depression and suicidality covariates. There were no subnetworks that were hyperconnected in individuals with high family risk of suicidality.

## 4. Discussion

To the best of our knowledge, this is the first study to investigate differences in white matter connectivity in individuals at high family risk of depression and high family risk of suicidality using graph theory methods. These methods allow for improved understanding of the network mechanisms underlying intergenerational psychiatric risk. We found evidence of hypoconnected subnetworks of subcortical-cortical regions associated with high family risk of MDD, and also with high family risk of suicidality, indicating disrupted information processing in these groups. This disrupted processing may be indicative of concurrent but not future anxiety and depression symptoms. Hypoconnected subnetworks associated with high family risk of depression differed from those associated with family risk of suicidality.

Graph summary measures indicated hypoconnectivity in individuals with high family risk of MDD. A lower clustering coefficient in individuals with a family history of MDD suggests that there are fewer connections between a region’s neighbors, also referred to as less segregation. Less segregation implies that these individuals can less efficiently communicate information within a given local brain area and are less able to process specialized functions in clustered subnetworks(32, 53, 54), suggesting that this may be a familial risk factor for developing depression. However, there was no consistent difference in global efficiency, or characteristic path length based on family history of MDD or on family history of suicidality. This suggests that MDD and suicidality family risk is not associated with network integration, and information transmission measured by characteristic pathlength and global efficiency(32, 53, 54). Thus, our findings show evidence of some disrupted processing in local subnetworks in people with high family risk of depression.

In previous studies on personal psychopathology, using functional connectivity networks, lower clustering coefficients and characteristic path length were present among individuals with MDD (55, 56) and lower clustering coefficients and efficiencies were present among individuals with a history of suicidality (35). In structural DTI networks, lower clustering coefficients and efficiencies were present among individuals with MDD (57, 58). However, findings and measures used vary by study, and some studies found no significant associations (34, 59, 60). In addition to focusing only on personal, not family history, the majority of these studies differed from ours by using functional networks(61), and most studies used self-reported symptoms as compared to the gold-standard diagnostic interviews we use here. However, these findings suggest similarities between individuals with personal MDD or suicidality and those at familial risk of disorder.

NBS analysis identified hypoconnected subnetworks in individuals at high family risk of MDD and suicidality, including regions previously associated with MDD and suicidality. The subnetwork for high family risk of depression centered on the left caudal middle frontal gyrus with hypoconnectivity to the contralateral inferior frontal gyrus, superior frontal gyrus, middle frontal gyrus and putamen. The middle frontal gyrus plays a role in attention, language processing, and working memory(62, 63), and its structural as well as functional connectivity has been associated with MDD(64, 65). This subnetwork also included the putamen, which plays a role in reward processing and anhedonia(66, 67). It has been associated with personal(56, 68–70) as well as family history(23, 25) of MDD through altered structure and function including decreased volume, increased aging, and aberrant activation. These regions are also part of the default mode network which is associated with self-reflection and rumination and shows aberrant functional connectivity in MDD(71–75). Subnetworks mostly included cross-hemispheric connections supporting the hypothesis that depression may be in part due to disrupted interhemispheric coordination(76–78).

Interestingly, a previous study investigating *functional* connectivity in an *a priori* defined fronto-temporo-parietal network using NBS in this cohort found familial risk of depression was associated with reduced influence of the inferior frontal gyri on information flow in the rest of the network(79), showing some overlap across functional and structural connectivity findings across different NBS analyses in these participants.

Hypoconnected subnetworks associated with high family risk of suicidality were largely symmetric, suggesting bilateral involvement and hypoconnectivity and included bilateral thalamus, bilateral precuneus, as well as bilateral superior frontal gyri and differed from those found for familial risk of MDD. Past NBS analysis has identified similar subnetworks associated with individual suicidality history including the rostral middle frontal gyrus, putamen, and thalamus(37–39, 80). Our findings are also in line with findings of decreased putamen, pallidum, and thalamus volume in individuals with a history of suicide attempts(30, 31). Adjusted models and removing individuals who had a personal history of suicidality, indicates hypoconnectivity beyond the effect of personal history of psychopathology or family history of MDD. The connections between the thalamus and the superior frontal gyrus associated with family risk of suicidality are part of the fronto-thalamic circuit, which has been suggested as a “suicide loop”(81, 82).

Strengths of this study include a unique multigenerational family study with diagnostic interviews of three generations of participants across multiple waves allowing for comprehensive understanding of suicidality and diagnostic history of depression as well as the usage of network analysis on this unique dataset. Limitations include that the population is primarily individuals of European descent from a limited geographical region, and additional studies are required to understand if these results are generalizable to a larger population. In addition, although comparable to other studies, the moderate sample size limits the ability to detect effects with small magnitudes.

Many studies have explored regions individually associated with personal history of MDD and personal history of suicidality, but little is known about the association between family history and white matter connectivity or brain network structure as a whole. We show that individuals with high family risk of MDD and high family risk of suicidality have brain-wide hypoconnectivity that suggests potential challenges in transfer of information and in processing specialized functions and was associated with increased anxiety and depressive symptoms.

Hypoconnectivity was concentrated in cortical-subcortical sub-networks that include the prefrontal cortex, putamen, thalamus, and precuneus. Altered brain connectivity associated with multigenerational risk goes beyond the effects of personal history and instead elucidates risk factors, emphasizing the clinical need for family history screening.

## Data Availability

Data analyzed in this study are from a subsample of the longitudinal Three Generations at High and Low Risk for Depression Study. In the last wave participants were approached to give consent for their data to be included in the NDA repository.
Data are being uploaded to the NDA for participants who provided consent, and will be made available to qualified investigators with the necessary approval.

## Acknowledgements

This project was supported by the National Institute of Mental Health *R01MH036197* (MMW, JP) and *K99MH129611* (MvD), an American Foundation for Suicide Prevention Awards *YIG-R-001-19* (MvD) and *SRG-0-130-16* (AT), and a Depression Center Pilot Award from the Columbia Department of Psychiatry (AT). The content is solely the responsibility of the authors and does not necessarily represent the official views of the National Institutes of Health, of American Foundation for Suicide Prevention or of any other sponsor.

