## SupplementalMaterials for "Differences in White Matter Structural Networks in Family Risk of Major Depressive Disorder and Suicidality: A Connectome Analysis"

Kelsall et al.

Supplemental Results

Below are additional adjusted model results for the NBS analyses. These models resulted from stepwise adjustment for family as well as individual history of suicidality and/or MDD, as detailed by the model description in Table 3.

Family risk of Depression Additional Models

After adjusting for personal history of MDD (Model 2), the subnetwork remained the same (p = 0.02579) except the putamen was not included in the subnetwork (Supplemental Figure 4) until a higher initial threshold of 1 x10^-3^.

After adjusting for personal history of suicidality (Model 3), the subnetwork was similar, however was only marginally significant at the threshold of 5x10^-4^ (p = 0.06079) and did not include the putamen or right rostral middle frontal gyrus until higher thresholds. Subnetworks were significant at higher thresholds. (Supplemental Figure 5)

After adjusting for family history of suicidality (Model 4), the subnetwork remained unchanged (p = 0.006799). (Supplemental Figure 6)

Family risk of suicidality Additional Models

After adjusting for personal history of suicidality (Model 2), two subnetworks remained connecting the left thalamus and the right superior frontal gyrus (p = 0.005799) and connecting right thalamus and the left superior frontal gyrus. (p = 0.03539) (Supplemental Figure 8)

After adjusting for personal history of MDD (Model 3), one subnetwork remained connecting the left thalamus and the right superior frontal gyrus. (p = 0.005199) (Supplemental Figure 9)

After adjusting for family history of MDD (Model 4), one subnetwork remained connecting the left thalamus and the right superior frontal gyrus. (p = 0.005399) (Supplemental Figure 10)

SUPPLEMENTAL TABLES AND FIGURES

Supplemental Table 1. All regions for Freesurfer whole brain segmentation.

| Cingulate | Caudal Anterior Cingulate |
| --- | --- |
|  | Isthmus Cingulate |
|  | Posterior Cingulate |
|  | Rostral Anterior Cingulate |
| Frontal | Caudal Middle Frontal Gyrus |
|  | Lateral Orbitofrontal Gyrus |
|  | Medial Orbitofrontal |
|  | Paracentral |
|  | Pars Opercularis |
|  | Pars Orbitalis |
|  | Pars Triangularis |
|  | Precental |
|  | Rostral Middle Frontal |
|  | Superior Frontal |
|  | Frontal Pole |
| Insula | Insula |
| Occipital | Cuneus |
|  | Lateral Occipital |
|  | Lingual |
|  | Pericalcarine |
| Parietal | Inferior Parietal |
|  | Postcentral |
|  | Precuneus |
|  | Superior Parietal |
|  | Supramarginal |

| Temporal | Banks of Superior Temporal Sulcus | |
| --- | --- | --- |
|  | Entorhinal Cortex | |
|  | Fusiform | |
|  | Inferior Temporal | |
|  | Middle Temporal | |
|  | Parahippocampal | |
|  | Superior Temporal | |
|  | Temporal Pole | |
|  | Transverse Temporal | |
| Subcortical | Cerebellum | |
|  | Thalamus | |
|  | Caudate | |
|  | Putamen | |
|  | Pallidum | |
|  | Amygdala | |
|  | Accumbens Area | |
|  | Hippocampal Subfields | Parasubiculum |
|  |  | Presubiculum |
|  |  | Subiculum |
|  |  | CA1 |
|  |  | CA2 |
|  |  | CA3 |
|  |  | CA4 |
|  |  | GC-DG |
|  |  | HATA |
|  |  | Fimbria |
|  |  | Molecular Layer |
|  |  | Hippocampal Fissure |
|  |  | Hippocampal Tail |

Supplemental Table 2. Standardized beta coefficients (SE) for GEE models comparing High Family Risk of MDD vs. Low Family Risk of MDD global and local graph measures. (^***^ p-value < .005, ^**^ p-value < .05)

|  | | Clustering Coefficient | | Characteristic Path Length | | Global Efficiency | | Local Efficiency | |
| --- | --- | --- | --- | --- | --- | --- | --- | --- | --- |
|  | | β ± SE | p value | β ± SE | p value | β ± SE | p value | β ± SE | p value |
| No Threshold | -- | **-0.377± 0.181** | 0.037 | -0.232  ±0.205 | 0.258 | 0.011  ±0.170 | 0.949 | -0.106  ±0.147 | 0.470 |
| Consistency Threshold | 50% | **-0.629± 0.168** | 0.00018 | -0.032  ±0.209 | 0.878 | 0.011  ±0.170 | 0.950 | **-0.398**  **±0.166** | **0.016** |
|  | 70% | **-0.495± 0.165** | 0.003 | -0.076  ±0.202 | 0.707 | 0.011  ±0.170 | 0.950 | -0.098  ±0.185 | 0.597 |
|  | 90% | **-0.909± 0.150** | < 0.00001 | -0.152  ±0.228 | 0.507 | 0.009  ±0.170 | 0.960 | -0.370  ±0.189 | 0.051 |

Supplemental Table 3. Standardized beta coefficients for GEE models comparing High Family Risk of MDD vs. Low Family Risk of MDD global and local graph measures after adjusting for personal history of MDD. (^***^ p-value < .005, ^**^ p-value < .05)

|  | | Clustering Coefficient | | Characteristic Path Length | | Global Efficiency | | Local Efficiency | |
| --- | --- | --- | --- | --- | --- | --- | --- | --- | --- |
|  | | β ± SE | p value | β ± SE | p value | β ± SE | p value | β ± SE | p value |
| No Threshold | -- | -0.286± 0.124 | 0.124 | -0.295± 0.221 | 0.183 | 0.118  ± 0.160 | 0.460 | -0.141  ± 0.144 | 0.329 |
| Consistency Threshold | 50% | **-0.526± 0.162** | **0.001** | -0.128  ± 0.224 | 0.569 | 0.118  ± 0.160 | 0.461 | **-0.461± 0.170** | **0.007** |
|  | 70% | **-0.374± 0.155** | **0.016** | -0.067  ± 0.217 | 0.758 | 0.118  ± 0.160 | 0.461 | -0.175  ± 0.189 | 0.354 |
|  | 90% | **-0.810± 0.136** | **< 0.00001** | -0.130  ± 0.201 | 0.518 | 0.116  ± 0.160 | 0.470 | **-0.471± 0.181** | **0.010** |

Supplemental Table 4. Standardized beta coefficients for GEE models comparing High Family Risk of MDD vs. Low Family Risk of MDD global and local graph measures after adjusting for personal history of suicidality. (^***^ p-value < .005, ^**^ p-value < .05)

|  | | Clustering Coefficient | | Characteristic Path Length | | Global Efficiency | | Local Efficiency | |
| --- | --- | --- | --- | --- | --- | --- | --- | --- | --- |
|  | | β ± SE | p value | β ± SE | p value | β ± SE | p value | β ± SE | p value |
| No Threshold | -- | -0.358  ±0.185 | 0.054 | -0.262  ± 0.216 | 0.224 | 0.049  ± 0.160 | 0.762 | -0.113  ± 0.149 | 0.447 |
| Consistency Threshold | 50% | **-0.553± 0.176** | **0.002** | -0.063  ± 0.211 | 0.767 | 0.049  ± 0.160 | 0.763 | **-0.324± 0.159** | **0.042** |
|  | 70% | **-0.476± 0.182** | **0.009** | -0.047  ± 0.205 | 0.817 | 0.049  ± 0.160 | 0.763 | -0.057  ± 0.177 | 0.746 |
|  | 90% | **-0.791± 0.176** | **0.00001** | -0.128  ± 0.223 | 0.567 | 0.1047  ± 0.161 | 0.772 | **-0.355± 0.185** | **0.055** |

Supplemental Table 5. Standardized beta coefficients for GEE models comparing High Family Risk of MDD vs. Low Family Risk of MDD global and local graph measures after adjusting for family history of suicidality. (^***^ p-value < .005, ^**^ p-value < .05)

|  | | Clustering Coefficient | | Characteristic Path Length | | Global Efficiency | | Local Efficiency | |
| --- | --- | --- | --- | --- | --- | --- | --- | --- | --- |
|  | | β ± SE | p value | β ± SE | p value | β ± SE | p value | β ± SE | p value |
| No Threshold | -- | **-0.363± 0.184** | **0.049** | -0.198  ± 0.212 | 0.351 | 0.040  ± 0.165 | 0.809 | -0.109  ± 0.145 | 0.474 |
| Consistency Threshold | 50% | **-0.547± 0.170** | **0.002** | -0.0002  ± 0.211 | 1.000 | 0.040  ± 0.165 | 0.809 | **-0.310**  **± 0.167** | **0.474** |
|  | 70% | **-0.461± 0.165** | **0.006** | -0.045  ± 0.198 | 0.821 | 0.040  ± 0.165 | 0.810 | -0.049  ± 0.187 | 0.794 |
|  | 90% | **-0.766± 0.153** | **0.00001** | -0.131  ± 0.222 | 0.557 | 0.038  ± 0.165 | 0.820 | -0.350  ± 0.190 | 0.066 |

Supplemental Table 6. Standardized beta coefficients for GEE models comparing High Family Risk of MDD vs. Low Family Risk of MDD global and local graph measures after adjusting for personal history of MDD, and personal and family history of suicidality. (^***^ p-value < .005, ^**^ p-value < .05)

|  | | Clustering Coefficient | | Characteristic Path Length | | Global Efficiency | | Local Efficiency | |
| --- | --- | --- | --- | --- | --- | --- | --- | --- | --- |
|  | | β ± SE | p value | β ± SE | p value | β ± SE | p value | β ± SE | p value |
| No Threshold | -- | -0.289  ± 0.186 | 0.121 | -0.266  ± 0.219 | 0.177 | 0.118  ± 0.157 | 0.450 | -0.146  ± 0.145 | 0.312 |
| Consistency Threshold | 50% | **-0.470± 0.167** | **0.005** | -0.122  ± 0.219 | 0.577 | 0.118  ± 0.157 | 0.451 | **-0.367± 0.168** | **0.029** |
|  | 70% | **-0.376± 0.163** | **0.021** | -0.052  ± 0.213 | 0.807 | 0.118  ± 0.157 | 0.451 | -0.114  ± 0.186 | 0.540 |
|  | 90% | **-0.703± 0.151** | **0.00001** | -0.124  ± 0.202 | 0.541 | 0.116  ± 0.157 | 0.461 | **-0.430± 0.181** | **0.0108** |

Supplemental Table 7. Standardized beta coefficients (SE) for GEE models comparing subjects at high and low family risk of suicidality using global and local graph measures. (^***^ p-value < .005, ^**^ p-value < .05)

|  | | Clustering Coefficient | | Characteristic Path Length | | Global Efficiency | | Local Efficiency | |
| --- | --- | --- | --- | --- | --- | --- | --- | --- | --- |
|  | | β ± SE | p value | β ± SE | p value | β ± SE | p value | β ± SE | p value |
| No Threshold | -- | -0.151  ±(0.214) | 0.481 | -0.313  ±0.253 | 0.217 | -0.279  ±0.160 | 0.081 | -0.246  ±0.154 | 0.111 |
| Consistency Threshold | 50% | -0.362  ±(0.211) | 0.087 | -0.097  ±0.264 | 0.715 | -0.280  ±0.160 | 0.081 | **-0.452± 0.160** | **0.005** |
|  | 70% | -0.231  ±(0.198) | 0.243 | -0.178  ±0.280 | 0.525 | -0.280  ±0.160 | 0.081 | 0.206  ±0.198 | 0.299 |
|  | 90% | **-1.452± (0.153)** | **< 0.00001** | -0.365  ±0.315 | 0.247 | -0.278  ±0.160 | 0.083 | -0.066  ±0.220 | 0.765 |

Supplemental Table 8. Standardized beta coefficients for GEE models comparing High Family risk of suicidality vs. Low Family risk of suicidality global and local graph measures after adjusting for personal history of suicidality. (^***^ p-value < .005, ^**^ p-value < .05)

|  | | Clustering Coefficient | | Characteristic Path Length | | Global Efficiency | | Local Efficiency | |
| --- | --- | --- | --- | --- | --- | --- | --- | --- | --- |
|  | | β ± SE | p value | β ± SE | p value | β ± SE | p value | β ± SE | p value |
| No Threshold | -- | -0.111  ± 0.231 | 0.633 | -0.389  ± 0.256 | 0.129 | -0.243  ± 0.158 | 0.123 | -0.249  ± 0.165 | 0.131 |
| Consistency Threshold | 50% | -0.273  ± 0.231 | 0.238 | -0.194  ± 0.273 | 0.477 | -0.244  ± 0.158 | 0.122 | **-0.412**  **± 0.165** | **0.013** |
|  | 70% | -0.212  ± 0.227 | 0.351 | -0.133  ± 0.290 | 0.647 | -0.244  ± 0.158 | 0.123 | 0.200  ± 0.200 | 0.319 |
|  | 90% | **-1.310± 0.194** | **< 0.0001** | -0.339  ± 0.323 | 0.294 | -0.242  ± 0.158 | 0.125 | -0.037  ± 0.225 | 0.871 |

Supplemental Table 9. Standardized beta coefficients for GEE models comparing high family risk of suicidality vs. low family risk of suicidality global and local graph measures after adjusting for personal history of mdd. (^***^ p-value < .005, ^**^ p-value < .05)

|  | | Clustering Coefficient | | Characteristic Path Length | | Global Efficiency | | Local Efficiency | |
| --- | --- | --- | --- | --- | --- | --- | --- | --- | --- |
|  | | β ± SE | p value | β ± SE | p value | β ± SE | p value | β ± SE | p value |
| No Threshold | -- | -0.011  ± 0.220 | 0.961 | -0.398  ± 0.280 | 0.155 | -0.183  ± 0.160 | 0.252 | **-0.304**  **± 0.149** | **0.042** |
| Consistency Threshold | 50% | -0.215  ± 0.208 | 0.301 | -0.202  ± 0.290 | 0.485 | -0.184  ± 0.160 | 0.250 | **-0.559**  **± 0.156** | **0.00036** |
|  | 70% | -0.076  ± 0.678 | 0.678 | -0.154  ± 0.301 | 0.609 | -0.184  ± 0.160 | 0.252 | 0.094  ± 0.200 | 0.640 |
|  | 90% | **-1.366**  **± 0.137** | **< 0.0001** | -0.380  ± 0.332 | 0.251 | -0.182  ± 0.160 | 0.256 | -0.175  ± 0.224 | 0.432 |

Supplemental Table 10. Standardized beta coefficients for GEE models comparing High Family risk of suicidality vs. Low Family risk of suicidality global and local graph measures after adjusting for Family History of MDD. (^***^ p-value < .005, ^**^ p-value < .05)

|  | | Clustering Coefficient | | Characteristic Path Length | | Global Efficiency | | Local Efficiency | |
| --- | --- | --- | --- | --- | --- | --- | --- | --- | --- |
|  | | β ± SE | p value | β ± SE | p value | β ± SE | p value | β ± SE | p value |
| No Threshold | -- | -0.106  ± 0.231 | 0.618 | -0.285  ± 0.261 | 0.274 | -0.282  ± 0.153 | 0.067 | -0.246  ± 0.151 | 0.105 |
| Consistency Threshold | 50% | -0.247  ± 0.230 | 0.230 | -0.082  ± 0.261 | 0.754 | -0.283  ± 0.153 | 0.066 | **-0.398**  **± 0.159** | **0.013** |
|  | 70% | -0.179  ± 0.351 | 0.351 | -0.160  ± 0.279 | 0.567 | -0.282  ± 0.153 | 0.066 | 0.197  ± 0.195 | 0.312 |
|  | 90% | **-1.285**  **± 0.157** | **< 0.0001** | -0.353  ± 0.310 | 0.256 | -0.280  ± 0.153 | 0.068 | -0.028  ± 0.218 | 0.899 |

Supplemental Table 11. Standardized beta coefficients for GEE models comparing High Family risk of suicidality vs. Low Family risk of suicidality global and local graph measures after adjusting for personal history of suicidality, and personal and family history of MDD. (^***^ p-value < .005, ^**^ p-value < .05)

|  | | Clustering Coefficient | | Characteristic Path Length | | Global Efficiency | | Local Efficiency | |
| --- | --- | --- | --- | --- | --- | --- | --- | --- | --- |
|  | | β ± SE | p value | β ± SE | p value | β ± SE | p value | β ± SE | p value |
| No Threshold | -- | -0.021  ± 0.223 | 0.927 | -0.429  ± 0.277 | 0.122 | -0.183  ± 0.158 | 0.248 | -0.290  ± 0.156 | 0.063 |
| Consistency Threshold | 50% | -0.169  ± 0.215 | 0.430 | -0.244  ± 0.288 | 0.397 | -0.183  ± 0.158 | 0.066 | **-0.474**  **± 0.161** | **0.004** |
|  | 70% | -0.090  ± 0.197 | 0.647 | -0.135  ± 0.300 | 0.653 | -0.183  ± 0.158 | 0.248 | 0.130  ± 0.200 | 0.516 |
|  | 90% | **-1.236**  **± 0.164** | **< 0.0001** | -0.360  ± 0.332 | 0.278 | -0.181  ± 0.158 | 0.252 | -0.110  ± 0.224 | 0.623 |

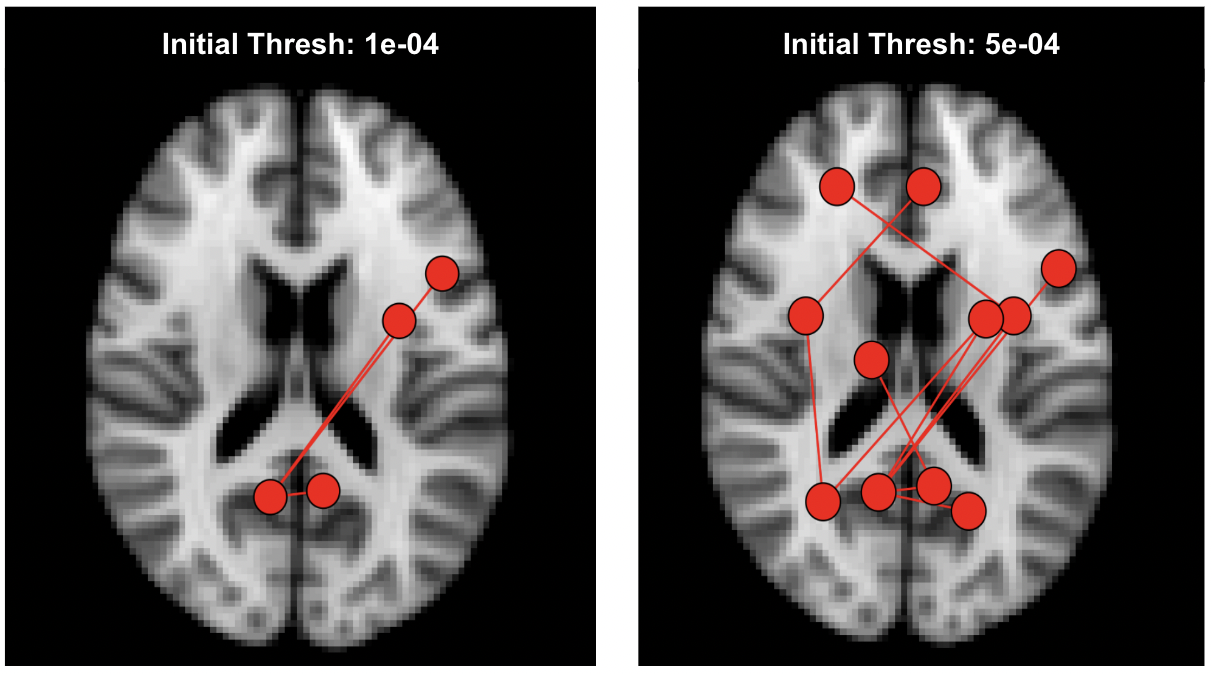

Supplemental Figure 1. Example comparison between two significant subnetworks based on different initial thresholds (p = 1x10^-4^ and p = 5x10^-4^) for the same set of subjects

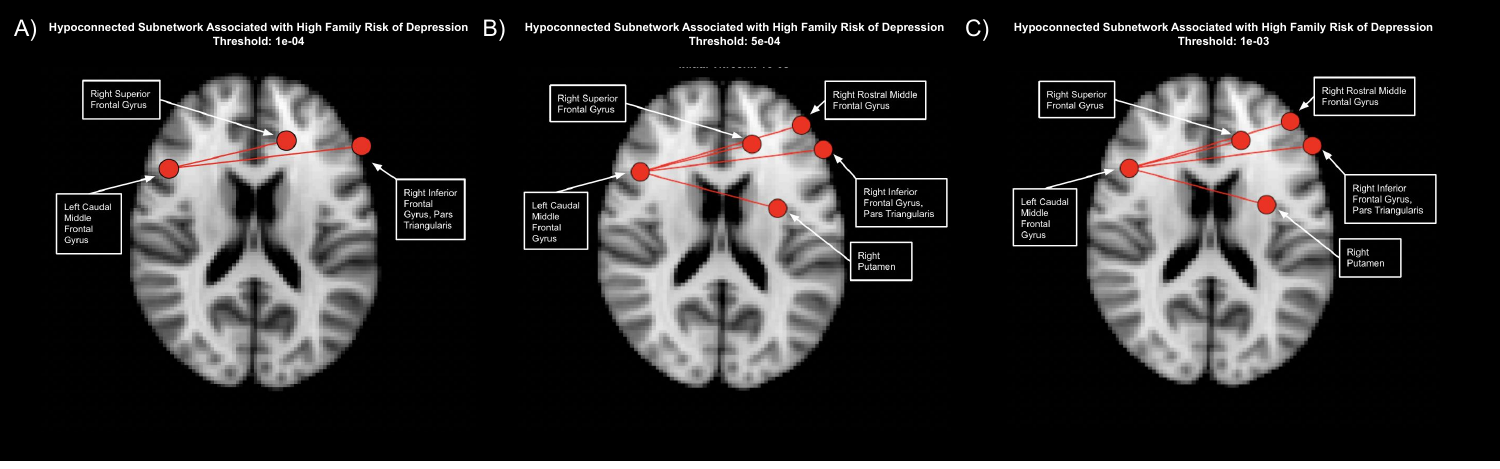

Supplemental Figure 2. Identified significant subnetworks with hypoconnectivity in high family risk of MDD subjects at multiple thresholds.  Subnetwork at A) 1e-04 (p = 0.008), B) 5e-04 (p = 0.0108, displayed in main analysis) and C) 1e-03 (p = 0.02498)

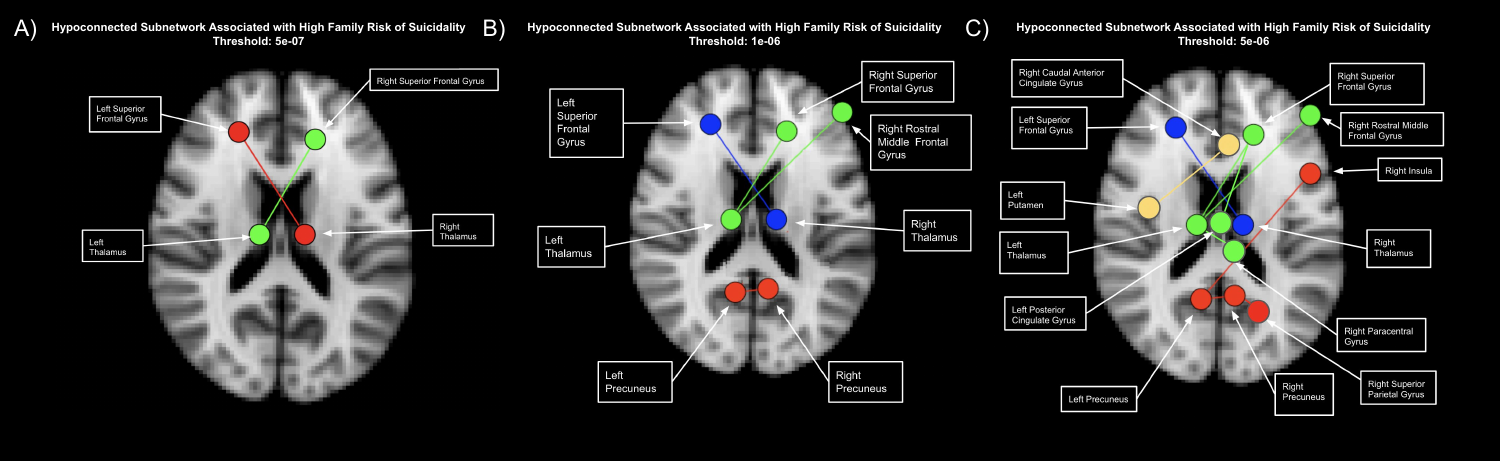

Supplemental Figure 3. Identified significant subnetworks with hypoconnectivity in high family risk of suicidality subjects at multiple thresholds.  Subnetwork at A) 5e-07 ( red network: p = 0.00040, and green network: p = 0.0046), B) 1e-06 (red network: p = 0.00040, blue network: p = 0.0046, and green network: p = 0.0046), displayed in main analysis) and C) 5e-06 (red network: p = 0.001, green network: p = 0.0002, blue network: p = 0.018, and yellow network: p = 0.018)

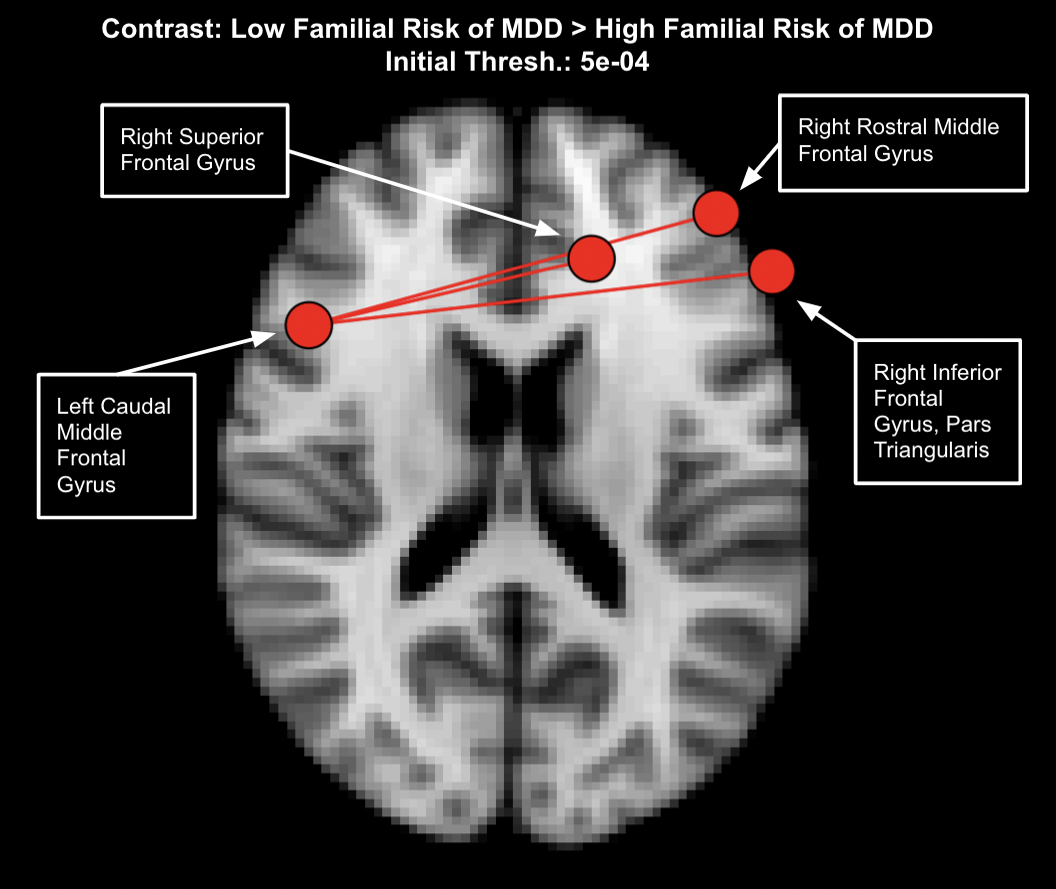

Supplemental Figure 4. Identified significant subnetwork with hypoconnectivity in high family risk of MDD subjects after adjusting for individual MDD. Subnetwork includes the four regions: left caudal middle frontal cortex, right superior frontal cortex, right rostral middle frontal cortex, and the right pars triangularis was significantly different (p = 0.02579) after permutation testing between the two risk groups.

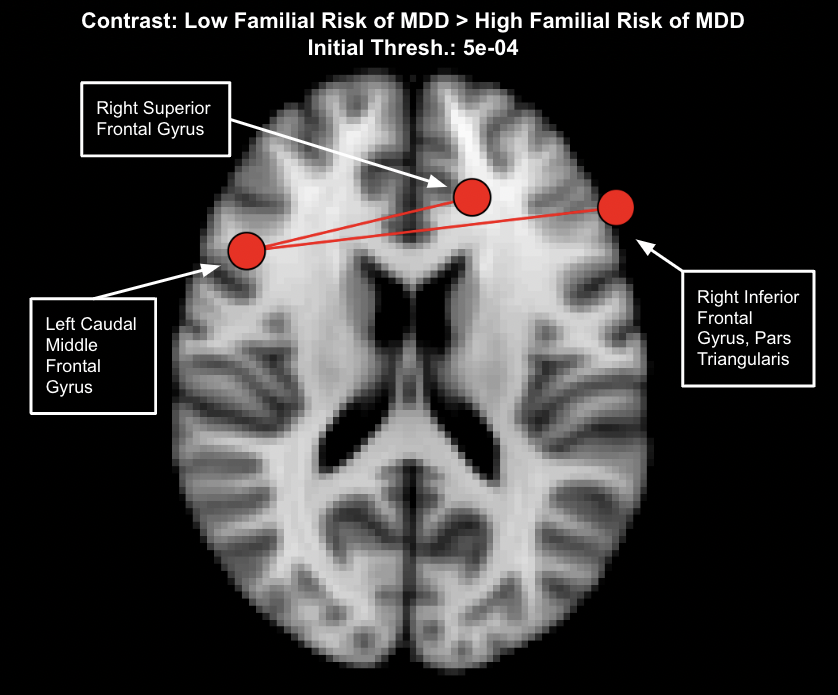

Supplemental Figure 5. Identified significant subnetwork with hypoconnectivity in high family risk of MDD subjects after adjusting for individual suicidality. Subnetwork includes the three regions: left caudal middle frontal cortex, right superior frontal cortex, and the right pars triangularis was only marginally significantly different (p = 0.06079) after permutation testing between the two risk groups.

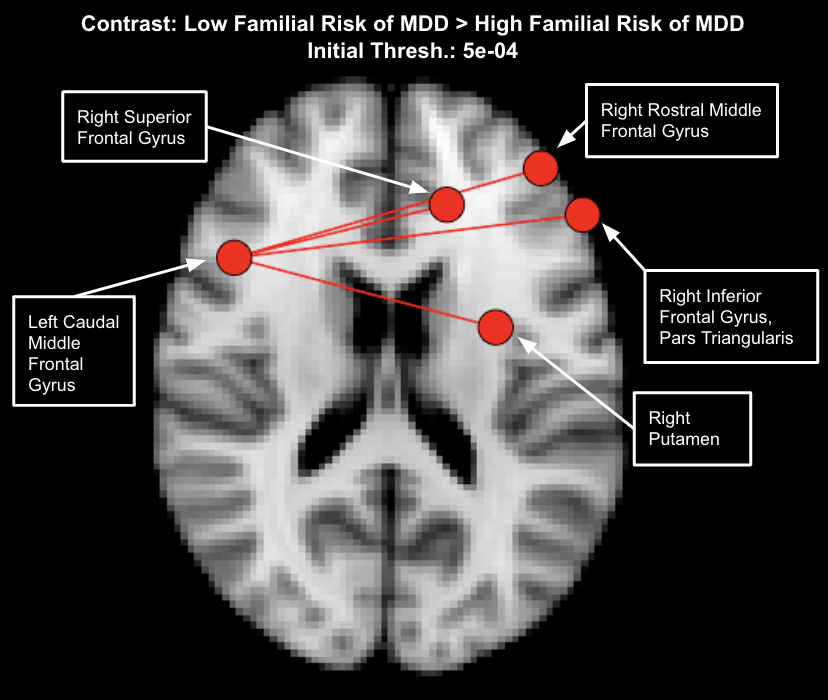

Supplemental Figure 6. Identified significant subnetwork with hypoconnectivity in high family risk of MDD subjects after adjusting for family history of suicidality. Subnetwork includes the four regions: left caudal middle frontal cortex, right superior frontal cortex, right rostral middle frontal cortex, and the right pars triangularis was significantly different (p = 0.006799) after permutation testing between the two risk groups.

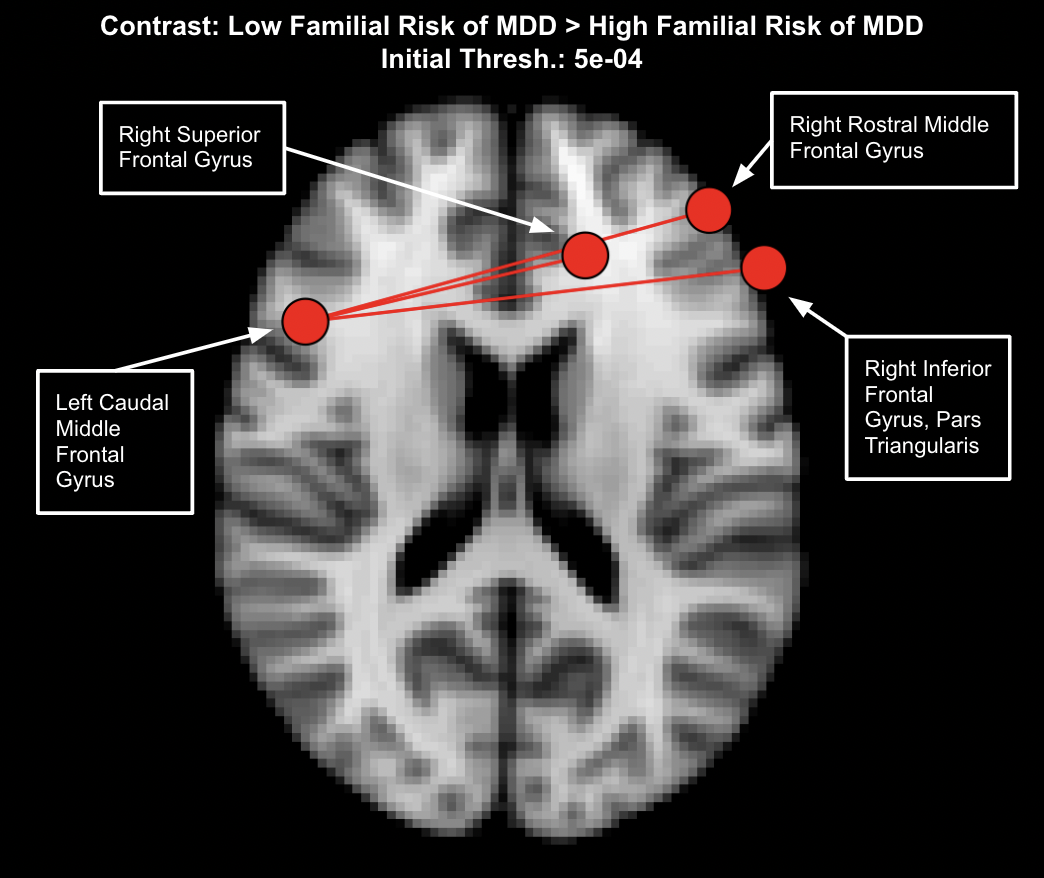

Supplemental Figure 7. Identified significant subnetwork with hypoconnectivity in high family risk of MDD subjects after adjusting for individual MDD, and family and individual history of suicidality. Subnetwork includes the four regions: left caudal middle frontal cortex, right superior frontal cortex, right rostral middle frontal cortex, and the right pars triangularis and was significantly different (p = 0.0212) after permutation testing between the two risk groups.

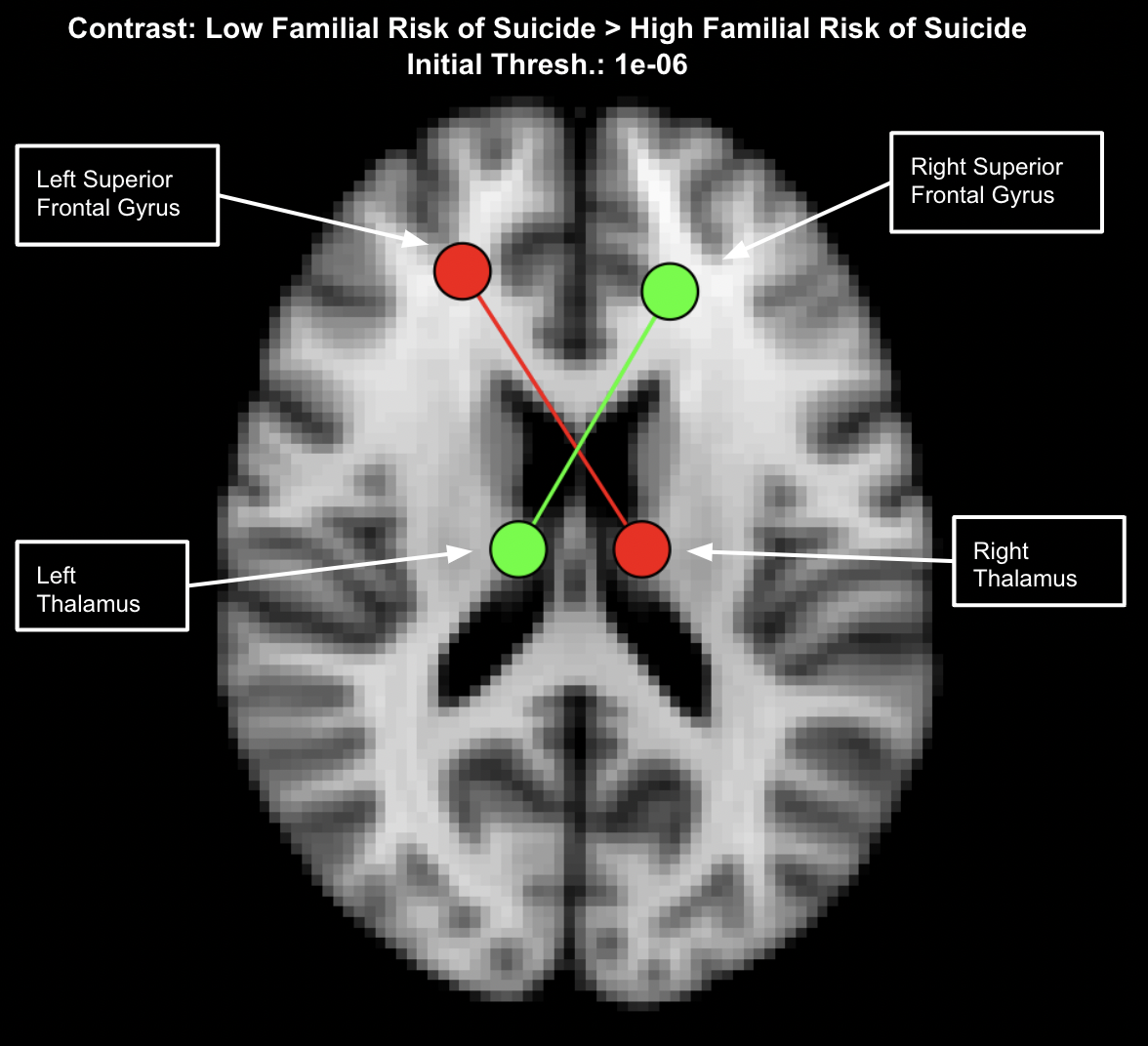

Supplemental Figure 8. Identified significant subnetwork with hypoconnectivity in high family risk of suicidality subjects after adjusting for individual suicidality. Subnetworks include left thalamus and the right superior frontal gyrus (p = 0.005799), and the right thalamus and the left superior frontal gyrus (p = 0.03539).

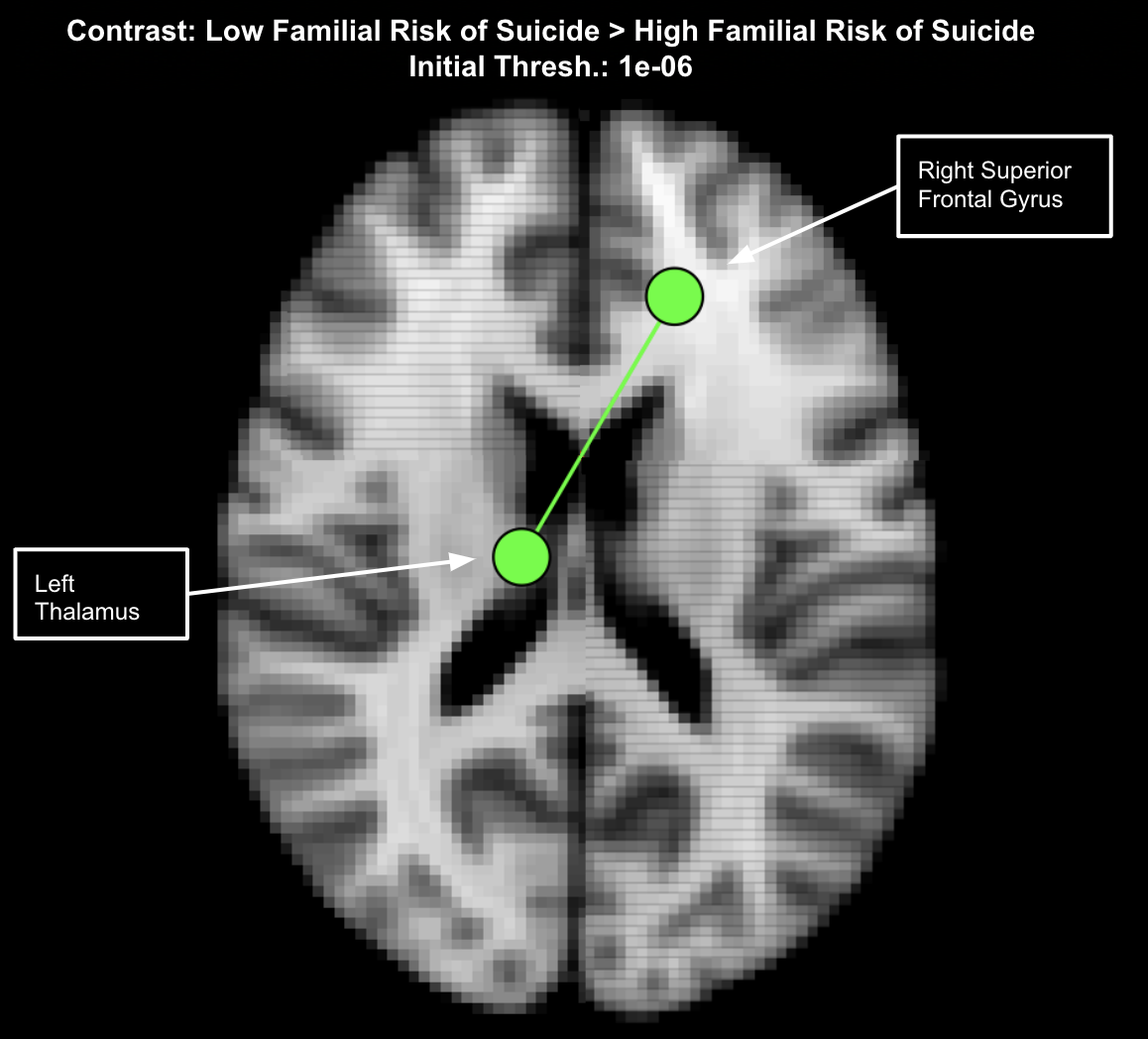

Supplemental Figure 9. Identified significant subnetwork with hypoconnectivity in high family risk of suicidality subjects after adjusting for individual MDD. Subnetwork includes left thalamus and the right superior frontal gyrus (p = 0.005199).

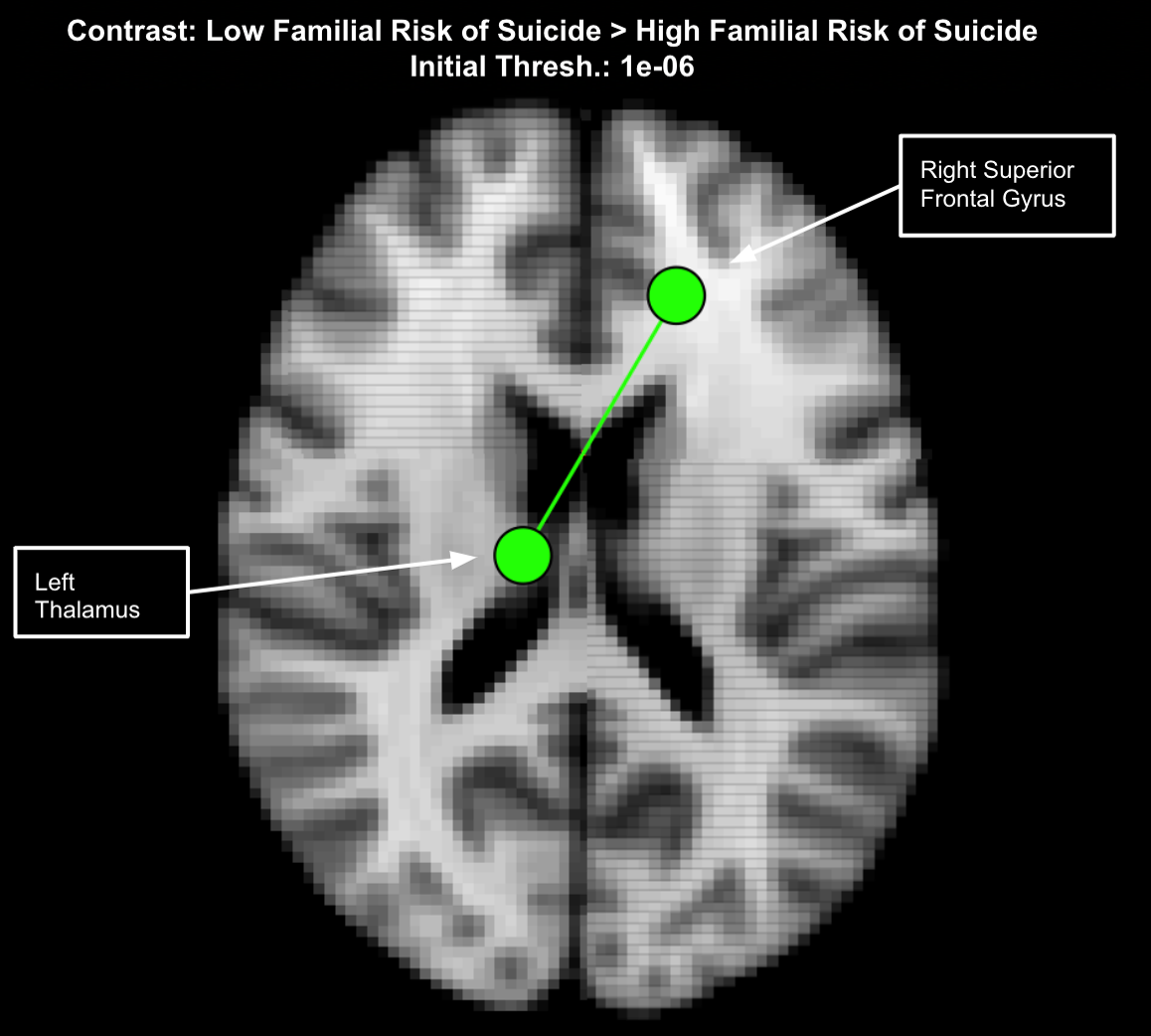

Supplemental Figure 10. Identified significant subnetwork with hypoconnectivity in high family risk of suicidality subjects after adjusting for family history of MDD. Subnetwork includes left thalamus and the right superior frontal gyrus (p = 0.005399).

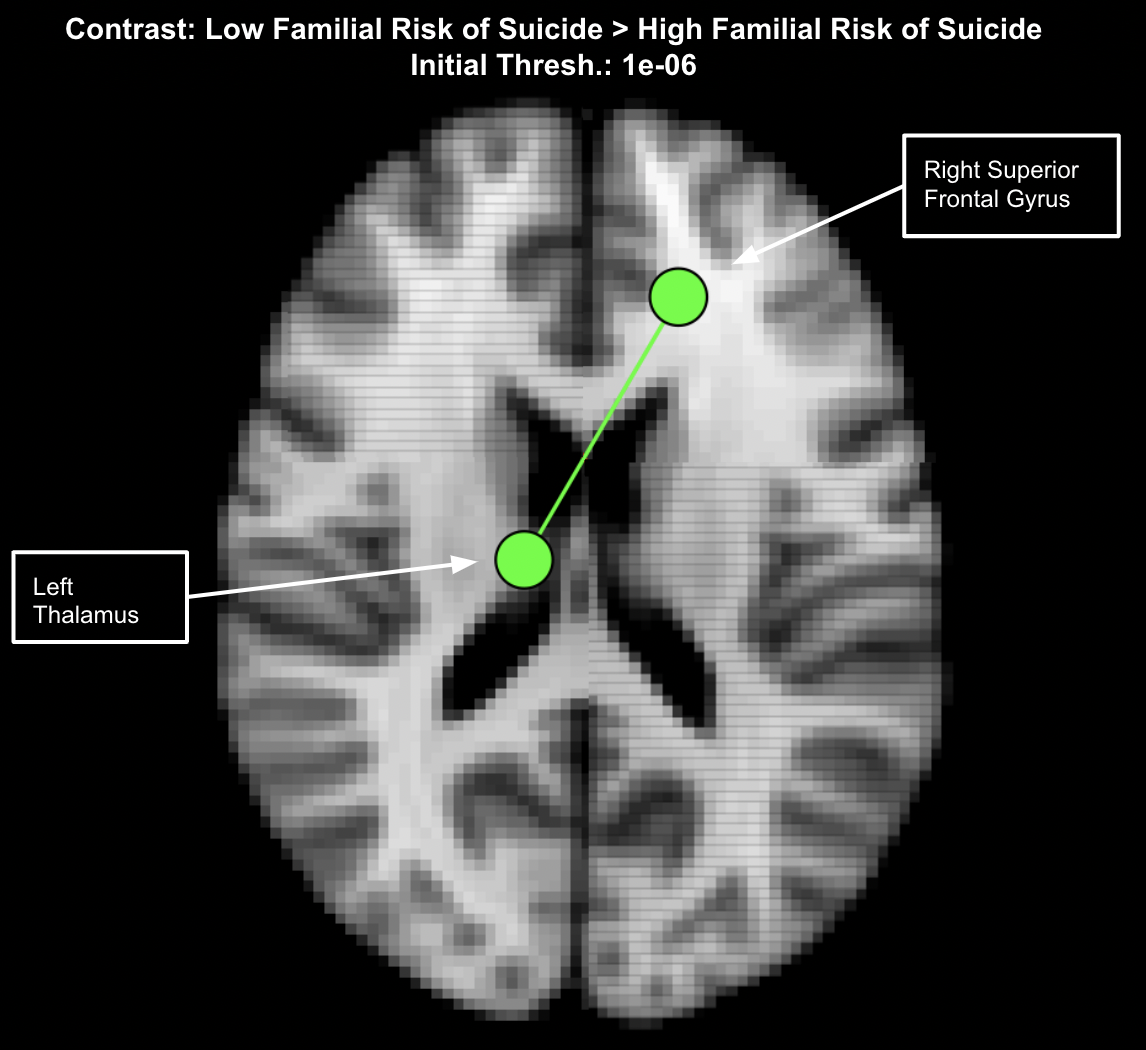

Supplemental Figure 11. Identified significant subnetwork with hypoconnectivity in high family risk of suicidality subjects after adjusting for family history of MDD, family history of suicidality and individual suicidality. Subnetwork includes left thalamus and the right superior frontal gyrus (p = 0.0024).

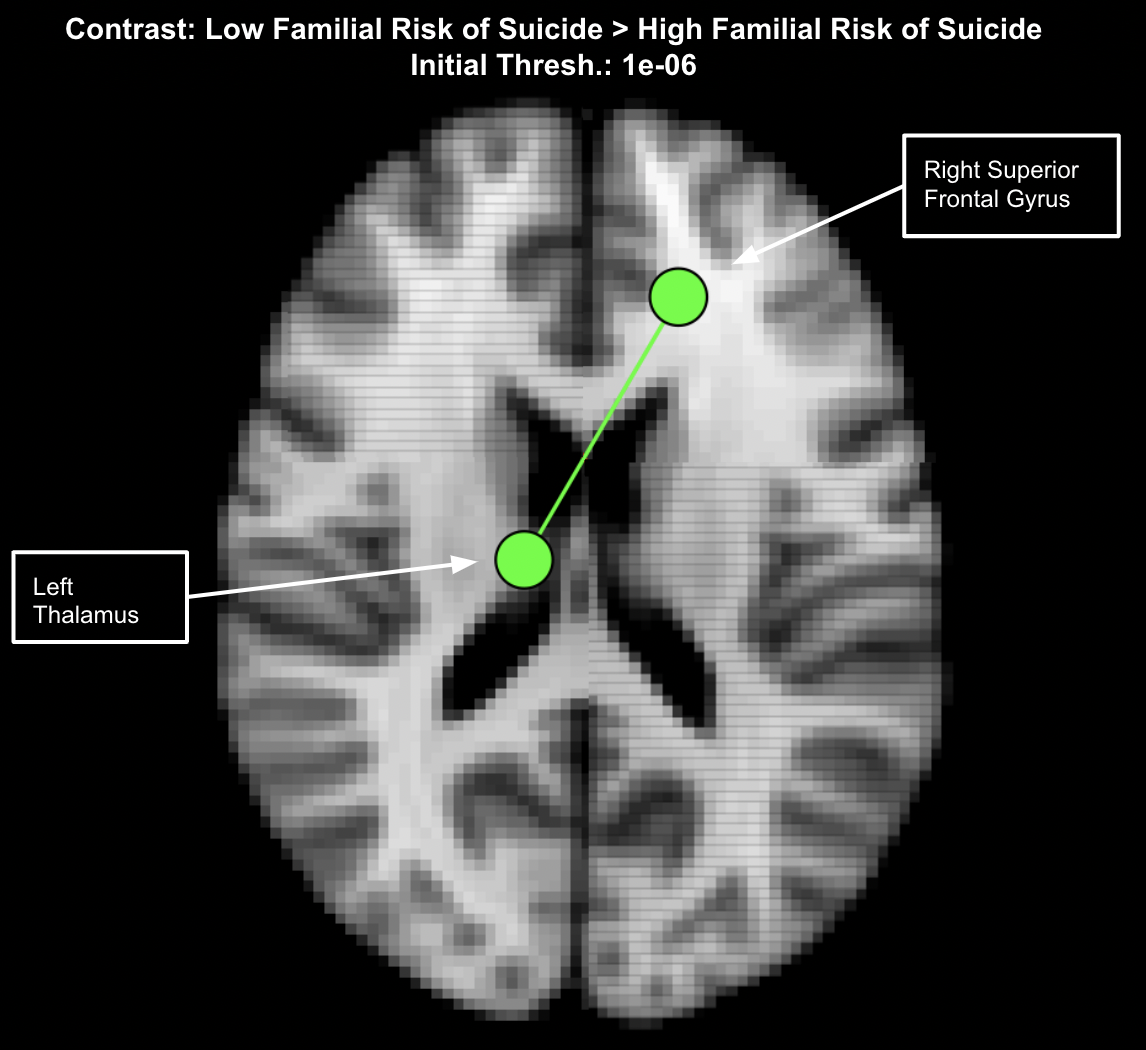

Supplemental Figure 12. Identified significant subnetwork with hypoconnectivity in high family risk of suicidality subjects after removing individuals with personal history of suicidality. Subnetwork includes left thalamus and the right superior frontal gyrus (p = 0.0014).
